# Evaluation of a Brief Educational Intervention to Improve Healthcare Provider Communication About Autism Genetic Testing

**DOI:** 10.64898/2026.11.19.26368524

**Authors:** Olivia G. Pilson, Lily S. Barna, Tyler Grimes, Taryn A. Salako, Gabriela M. Ramírez-Renta, Amy Wang, Chris Gunter

**Affiliations:** Engagement Methods Unit, Social and Behavioral Research Branch, National Human Genome Research Institute, Bethesda, MD; Northwestern University School of Communications, Evanston, IL; The EMMES Corporation, Rockville, MD; Columbia University Mailman School of Public Health, New York City, NY; University of Colorado Anschutz Medical School, Aurora, CO; Stony Brook University, Stony Brook, NY; Office of the Director, National Human Genome Research Institute, Bethesda, MD

**Keywords:** Autism, Neurodevelopmental, Genetic Testing, Educational Intervention, Healthcare Communication

## Abstract

Although professional guidelines recommend offering genetic testing following an autism diagnosis, uptake remains low in the United States. Healthcare providers frequently report limited education and uncertainty discussing autism genetics with patients and families. We developed and evaluated a brief educational intervention to improve provider communication about autism genetic testing, consisting of animated videos addressing autism genetics, available testing options, and strategies for discussing testing with autistic individuals and families. We evaluated the intervention among 100 U.S.-based healthcare providers and a community panel of 99 autistic adults. Participants completed a validated measure of genetic literacy before viewing the videos, then evaluated the intervention and its perceived impact on genetic testing. Providers showed higher genetic literacy than the general U.S. population, yet only 23% reported frequently discussing genetic testing before the intervention. Afterward, 86% reported greater likelihood of recommending genetic testing following diagnosis, and 88% reported the videos changed how they would discuss genetics with patients and families. Both cohorts rated the resources favorably, and 71% of autistic adults reported greater likelihood of pursuing genetic testing after viewing the videos. Brief, communication-focused educational resources may improve workflows for autism genetic testing and support informed decision-making for providers, autistic individuals, and their families.

## INTRODUCTION

The clinical use of genetic testing has expanded rapidly, increasing expectations that healthcare providers (HCPs) can appropriately recommend testing and communicate genetic information to patients and families. In 2020, the National Human Genome Research Institute made the “bold prediction” that genomic information would transition from “boutique to mainstream in all clinical settings” by 2030 (Green et al., 2020). Genetic literacy and provider training in both genetics and patient-centered communication are crucial foundations for this goal, particularly in conditions where genetic and environmental factors may play a role (Ramírez Renta et al., 2025). Autism is a neurodevelopmental condition with a substantial genetic contribution and marked phenotypic heterogeneity (Olson et al., 2024). Although genetic testing does not establish an autism diagnosis, professional guidelines recommend offering genetic testing as part of the clinical evaluation to identify potential etiologic factors and associated medical risks (Hyman et al., 2020; Schaefer & Mendelsohn, 2013). Nevertheless, uptake of genetic testing among autistic individuals remains low, indicating persistent gaps between clinical recommendations and practice (Arcebido et al., 2025; Brown et al., 2025). Provider specialty also influences both frequency and type of testing ordered, with non-genetics providers insufficiently enabled to order or promote referrals (Harrington et al., 2024). Prior work from our unit and others has identified provider-level barriers to recommending genetic testing for autism, including limited genetics education, uncertainty regarding clinical utility, and challenges in discussing testing with individuals and families (Abreu et al., 2022; Barton et al., 2017; Wang et al., 2024). For medical geneticists, this means that referrals typically arrive only after a separate, non-referring provider, often a psychologist or nurse practitioner, has already completed the diagnostic evaluation, adding a handoff step that can delay or prevent a patient from ever reaching a genetics clinic (Abreu et al., 2022; Wang et al., 2024).

Educational interventions, particularly videos and/or animations targeted to the providers involved at each stage of this pathway, represent a potential mechanism for addressing these communication and workflow barriers, an approach increasingly used both to prepare patients for genetic counseling in other specialties (Schneider et al., 2025) and, more recently, to streamline the referral-to-testing pathway within autism-specific genetics clinics themselves (Barton et al., 2025). In genomic medicine, brief digital and multimedia educational resources, including video-based materials, have been used to support knowledge acquisition and communication skills in scalable formats (Chen et al., 2024; Cutica et al., 2024; Daniel et al., 2025; Lewis et al., 2020). Not only can they be used as part of “just-in-time” genomic education for clinicians (Hoffman et al., 2026), but video-based modules also specifically have demonstrated educational value on the parent and family side of autism genetic testing information (Xu et al., 2018).

In this study, we developed a three-part video-based educational intervention addressing (1) foundational concepts in autism genetics, (2) types and potential benefits of genetic testing, and (3) approaches to discussing genetic testing with individuals and families. Our objective was to evaluate both the perceived utility of these educational resources and their potential to improve communication about autism genetic testing by assessing healthcare providers’ self-reported intent to recommend testing and participants’ evaluations of the resources. We recognize the sensitivity around genetics language and testing in the autism community, which too often uses ableist ideas and terminology (Sugden et al., 2025). Therefore, we sought feedback from an autistic community panel to assess the clarity and appropriateness of the communication content, serving as a secondary evaluation of suitability rather than an efficacy outcome.

## METHODS

### Editorial Policies and Ethical Considerations

Two research protocols were deemed exempt by the Office of Research Protection of the National Institutes of Health (NIH) and approved by the Institutional Review Board at the National Institutes of Health (IRB001103 for the pilot, IRB002400 for larger study). All participants were given a summary of the study and consented to the study prior to administration of any intervention or survey.

### Development of Video Resources

From our previous study (Wang et al., 2024), we compiled areas that were flagged for developing educational resources for HCPs and then divided those resources into three sections as outlined above. Once we identified which topics were to be covered in each of the resources we then decided on additional examples and research to include to make the resources engaging. These concepts were included in our resource scripts. We then illustrated each of these areas in separate animated videos with narration using Adobe Animate and Adobe Premiere Pro and published them online using Vimeo (links in Supplemental Table 1).

### Pilot Study

First, we conducted a pilot study (Providing Autism and Genetics Education Study or PAGES I) gathering two rounds of feedback on the three educational intervention resources from a convenience sample (*n* = 5) healthcare professionals who have experience diagnosing autism. Both cycles of pilot study participants were contacted via email for their known involvement in diagnosing autism and history of recommending genetic testing. Pilot study participants were sent a link to the survey via SurveyMonkey after consenting to be a part of the study and were sent a $50 Amazon gift card via email upon completion.

The initial model for the pilot study was adapted from the Plan-Do-Study-Act (PDSA) Cycle for developing and assessing the quality of resources and activities in healthcare spaces (Taylor et al., 2014). Adapting the framework outline from Crowl et al. (Crowl et al., 2015) we requested quantitative feedback from pilot study participants in quantitative Likert scale ratings (4-point Likert scale) and qualitative open-ended input. Quantitative questions asked during the pilot (and later) study to gather feedback on resources were (1) How relevant did you find the information in this video for your clinical work? (2) How useful did you find the video to be for your clinical work? and (3) How likely are you to recommend this video to a colleague? We also requested qualitative feedback during the pilot study in the form of open text boxes, to gain further insight into how participants evaluated the resources. We incorporated the quantitative and qualitative feedback by modifying resource scripts and videos for future use.

### Adaptations to Resources and Survey Instrument

We made modifications and additions before administering the survey more widely: all quantitative Likert scale questions were changed from a 4-point to a 7-point Likert scale both to include a neutral option and to reflect the benefits and validity of 7-point Likert scale use in social research (Sullivan & Artino, 2013). More post-intervention evaluation questions about attitudes toward recommending genetic testing for autism were added at the end of the study. We also added in a 46-question genetic literacy assessment (Barna et al., 2026) before participants viewed the videos. Finally, based on pilot study participant feedback and community preferences, we adapted language and content included in the resources. Transcripts from the final videos are provided in the supplementary materials (Supplemental Document 1).

### Participants and Data Collection Measures

We conducted the full study (PAGES II) entirely online; it was designed to have a completion time of 30-45 minutes. The survey and participant answers were hosted on SurveyMonkey, this time reached through the platform Prolific, used to recruit prospective participants for the study. All participant information was kept anonymous by the platform; researchers only accessed and analyzed deidentified participant survey responses. All verified participants who completed the survey were provided with a $10 credit through Prolific.

First, we used the participant screening information on Prolific to recruit a random sample of 100 HCPs, who were eligible to participate if they had indicated that they were either nurses, psychologists, or doctors, are based in the United States and are comfortable reading and writing in English (approximately 900 eligible participants in the Prolific platform at time of data collection). Further questions about demographics and their healthcare specialty and involvement in the autism diagnostic process were presented after participants consented to being in the study. Second, we recruited a random sample of 100 autistic adults including participants who reported being diagnosed with autism (any age), were in the process of receiving a diagnosis, or identify as being autistic in Prolific (approximately 4,800 eligible participants). We later removed one participant from the autistic adult cohort (AC) as they had not completed the study. The final sample sizes for this study were 100 US-based HCPs and 99 US-based autistic adults. The respondents completed the survey in 30-55 minutes. Responses from the HCPs were collected in February 2025, and responses from the AC were collected in March 2025.

The content of the intervention for the HCPs surveyed and resources assessed by the panel of autistic adults was the same. However, we tailored the post-intervention questionnaire to each participant group so that the language of the questions would be more applicable to the surveyed group (see Supplemental Document 2 for HCPs and 3 for AC surveys). Finally, we gave participants the long version of the Education and Assessment of Genetic Literacy (EAGL-long) measure (Barna et al., 2026).

### Language Preferences

The phrases used to characterize autism are personal and should be based on how participants prefer to express themselves. Our unit, in previous research, has used the terms “autism,” “autism spectrum disorder,” or “ASD” to denote the condition and “autistic” to refer to individuals participating in this study. We now only use “autism” and “autistic” to acknowledge that language is continuously evolving and that community-favored guidelines for discussing autism-related information may continue to diverge and evolve (Bottema-Beutel et al., 2020; Sugden et al., 2025).

### Statistical Analysis

Reports and results from the pilot study were analyzed and kept separate from the rest of the study because of alterations and additions to the new survey instrument (PAGES II). We performed data preparation in Microsoft Excel. R Statistical version 4.5.3 (R Core Team, 2024), was used to generate descriptive statistics on participant demographics (education, age, etc.), genetic literacy subscale score, and quantitative resource evaluations (with visualization). We performed data analysis including Welch t-test comparisons with readxl (Wickham, 2025) and dplyr (Wickham, 2026). Additionally, we compared the scores between each of these groups to the published genetic literacy scores of a general population sample (Barna et al., 2026). We conducted exploratory Spearman rank correlation analyses to evaluate associations between recency of genetics/genomics continuing medical education and genetic literacy and adjusted for multiple comparisons using the Benjamini-Hochberg false discovery rate method.

### Analysis of Open-Ended Responses

Participants in both cohorts were invited to provide optional open-ended comments throughout the survey, serving different purposes between the two cohorts (Supplemental Document 1). Among HCPs, the questions elicited feedback on the content and clinical utility of the educational resources. Among autistic participants, they explored perspectives on autism genetic testing and whether the educational resources influenced those perspectives. Responses were reviewed descriptively to identify recurring perspectives on the educational resources and autism genetic testing. Because the purpose of these data was to contextualize the quantitative findings rather than perform a formal qualitative analysis, responses were not coded using a qualitative framework. Representative quotations were selected to illustrate recurring viewpoints based on the clarity and specificity of participants’ comments. All quotations were de-identified before inclusion. For HCP participants, healthcare specialty and involvement in the autism diagnostic process are reported to provide clinical context.

## RESULTS

### Pilot Study Results

The pilot study included five healthcare providers involved in autism diagnosis, comprising three psychologists in the first cycle and one psychologist and one developmental-behavioral pediatrician in the second cycle. All participants had diagnosed an autistic patient within the preceding six months. On a 7-point scale, participants reported moderate confidence in their genetic literacy (mean 3.8) and greater familiarity with autism genetics research (mean 5.6).

Participants evaluated each educational resource for relevance, usefulness, and likelihood of recommending it to a colleague using a 4-point Likert scale. Across all three domains, the third video, which focused on common questions patients and families ask about genetic testing, received the highest ratings, with mean scores of 3.6 for relevance, 3.6 for usefulness, and 3.2 for likelihood of recommendation. Quantitative ratings and qualitative feedback from the pilot informed revisions to the scripts, survey instrument, and final video resources before the full study.

### Participant Characteristics

The study included 100 U.S.-based healthcare providers (HCPs) and 99 U.S.-based autistic adults (AC). Demographic characteristics are summarized in Table 1. Based on Rural–Urban Continuum Codes (RUCCs) derived from participant ZIP codes, 85.9% of HCPs and 93.9% of AC participants resided in metropolitan areas. One HCP reported an invalid ZIP code and was excluded from this analysis. The metropolitan distribution of the HCP cohort closely matched that of the U.S. population, for which 86.2% of individuals reside in metropolitan counties according to the 2020 U.S. Census [https://www.ers.usda.gov/data-products/rural-urban-continuum-codes/documentation].

**Table 1:**
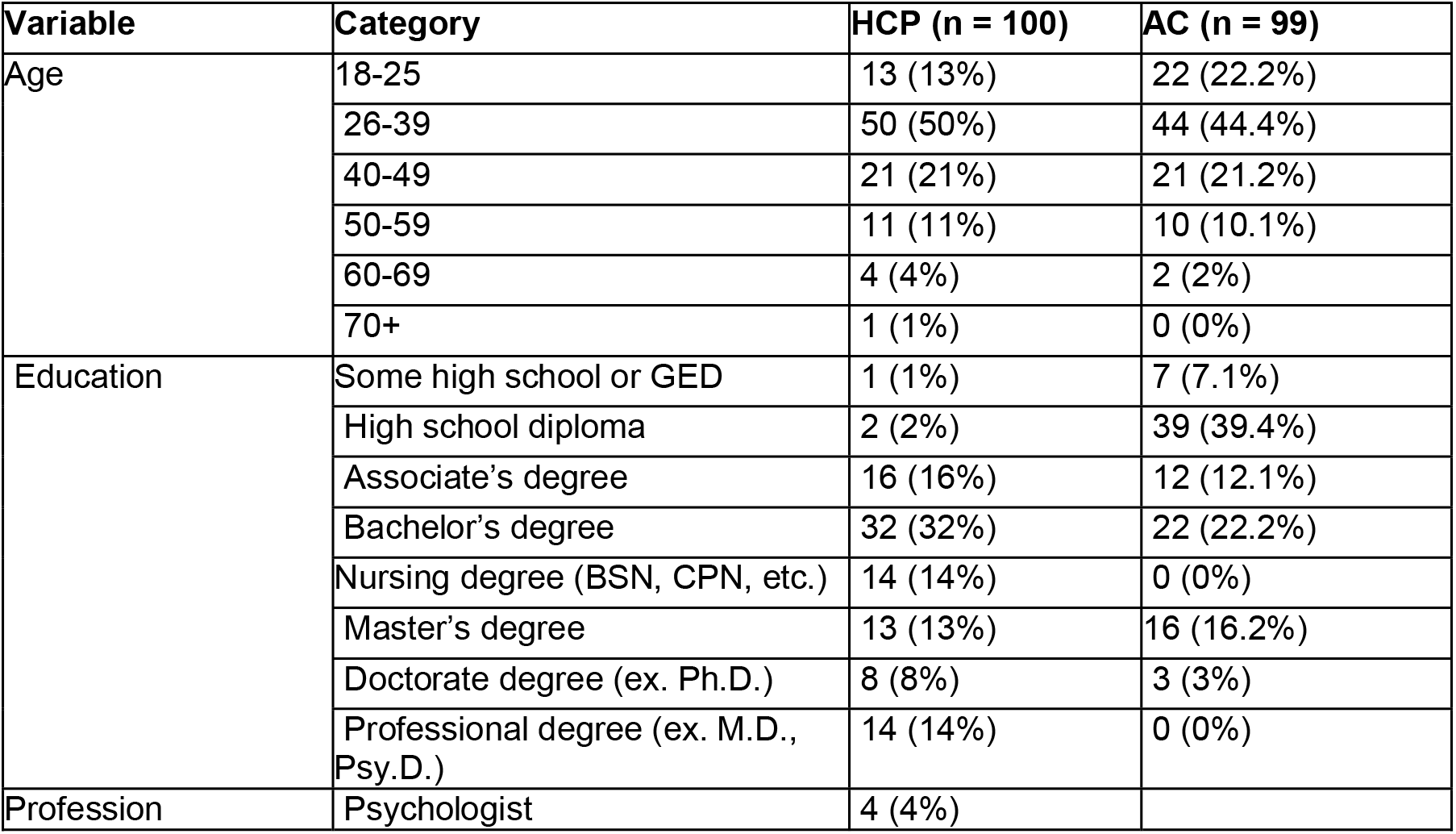
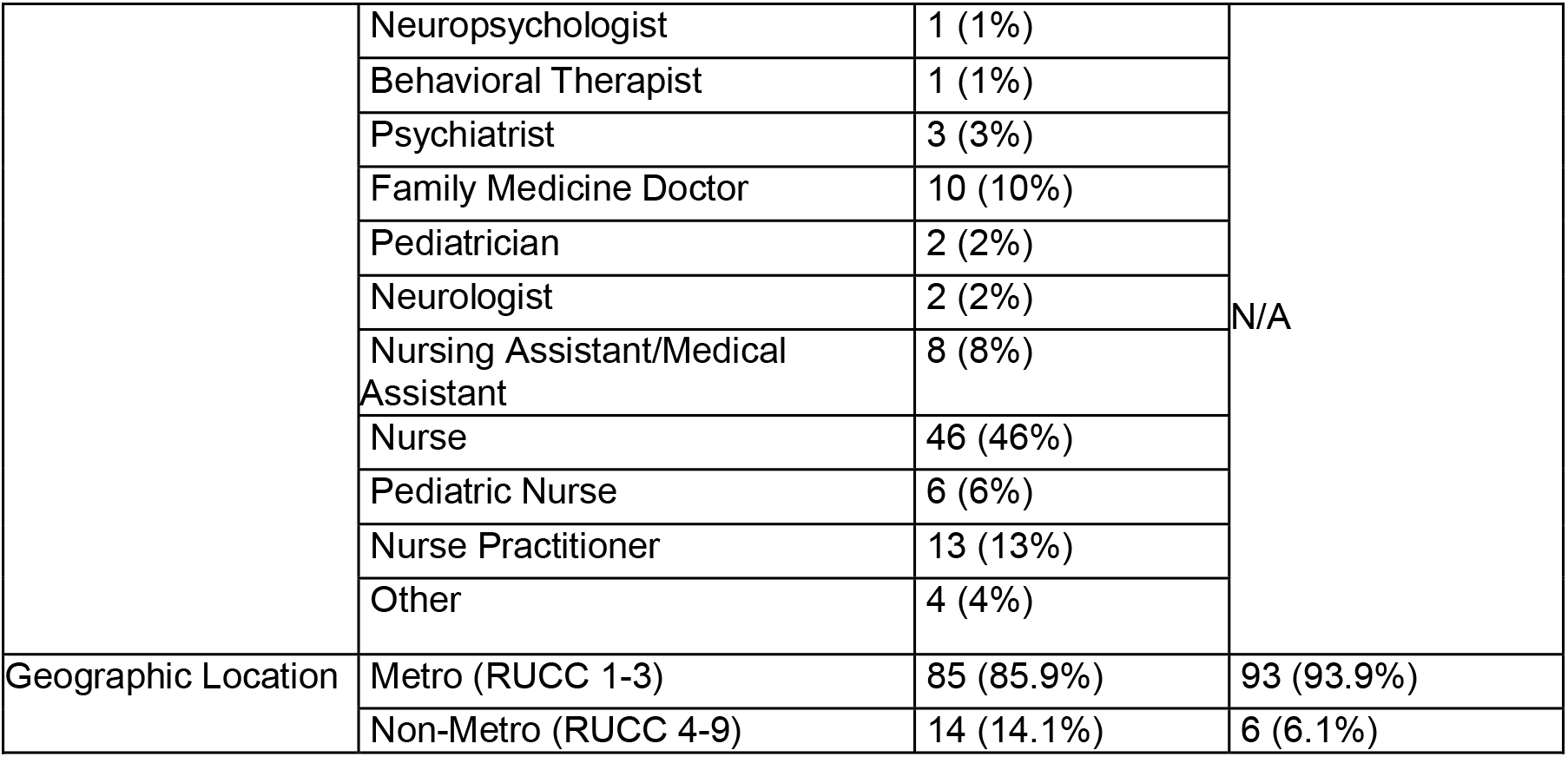
Reported demographics from HCP and autistic adults (AC)

| Variable | Category | HCP (n = 100) | AC (n = 99) |
|---|---|---|---|
| Age | 18-25 | 13 (13%) | 22 (22.2%) |
|  | 26-39 | 50 (50%) | 44 (44.4%) |
|  | 40-49 | 21 (21%) | 21 (21.2%) |
|  | 50-59 | 11 (11%) | 10 (10.1%) |
|  | 60-69 | 4 (4%) | 2 (2%) |
|  | 70+ | 1 (1%) | 0 (0%) |
| Education | Some high school or GED | 1 (1%) | 7 (7.1%) |
|  | High school diploma | 2 (2%) | 39 (39.4%) |
|  | Associate’s degree | 16 (16%) | 12 (12.1%) |
|  | Bachelor’s degree | 32 (32%) | 22 (22.2%) |
|  | Nursing degree (BSN, CPN, etc.) | 14 (14%) | 0 (0%) |
|  | Master’s degree | 13 (13%) | 16 (16.2%) |
|  | Doctorate degree (ex. Ph.D.) | 8 (8%) | 3 (3%) |
|  | Professional degree (ex. M.D., Psy.D.) | 14 (14%) | 0 (0%) |
| Profession | Psychologist | 4 (4%) |  |
|  | Neuropsychologist | 1 (1%) | N/A |
|  | Behavioral Therapist | 1 (1%) |  |
|  | Psychiatrist | 3 (3%) |  |
|  | Family Medicine Doctor | 10 (10%) |  |
|  | Pediatrician | 2 (2%) |  |
|  | Neurologist | 2 (2%) |  |
|  | Nursing Assistant/Medical Assistant | 8 (8%) |  |
|  | Nurse | 46 (46%) |  |
|  | Pediatric Nurse | 6 (6%) |  |
|  | Nurse Practitioner | 13 (13%) |  |
|  | Other | 4 (4%) |  |
| Geographic Location | Metro (RUCC 1-3) | 85 (85.9%) | 93 (93.9%) |
|  | Non-Metro (RUCC 4-9) | 14 (14.1%) | 6 (6.1%) |

Background characteristics relevant to autism genetics are summarized in Tables 2 and 3. Among HCPs, 33% reported completing genetics or genomics continuing medical education (CME) within the previous 1–5 years, whereas an additional 33% reported that their most recent genetics-focused CME had occurred more than five years earlier or that they had never completed genetics-focused CME. Sixty-seven HCPs reported involvement in the autism diagnostic process, including 20% who had diagnosed an autistic patient within the preceding six months. Although 75% reported discussing autism-related genetic testing with patients or families at least occasionally, only 23% reported doing so frequently or always, whereas 41% reported discussing it never or rarely. Finally, 39% of the HCP sample reported that either they or an immediate family member is autistic.

**Table 2:**
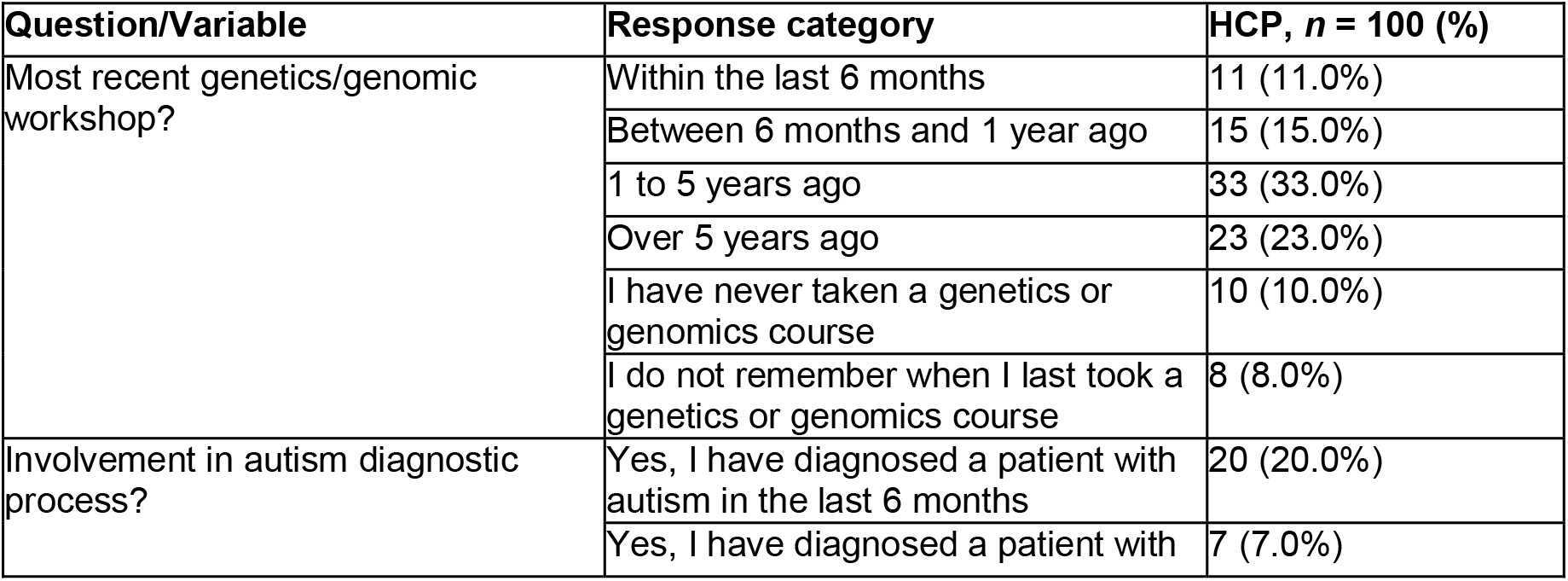
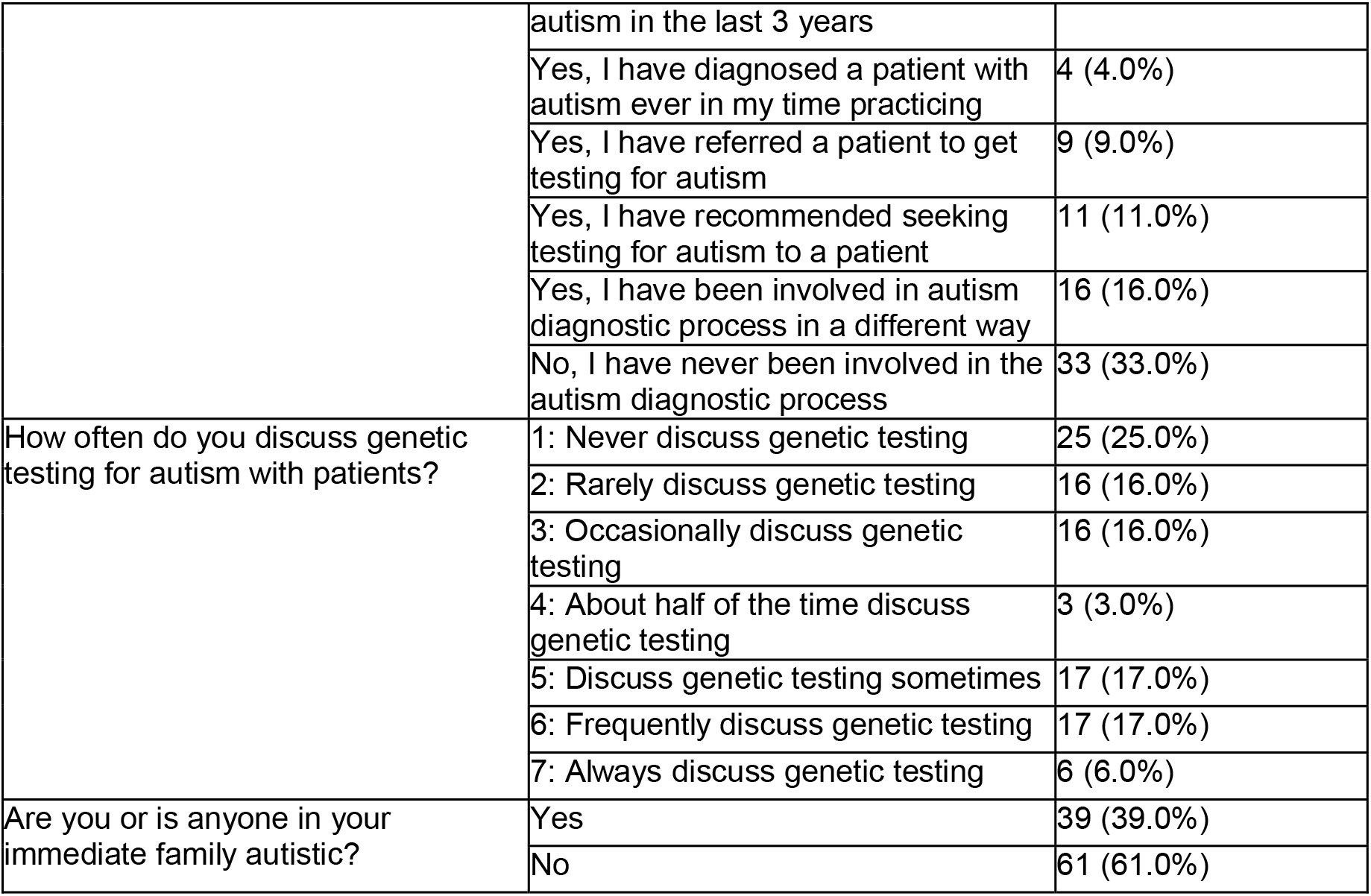
HCPs additional background questions.

**Table 3:**
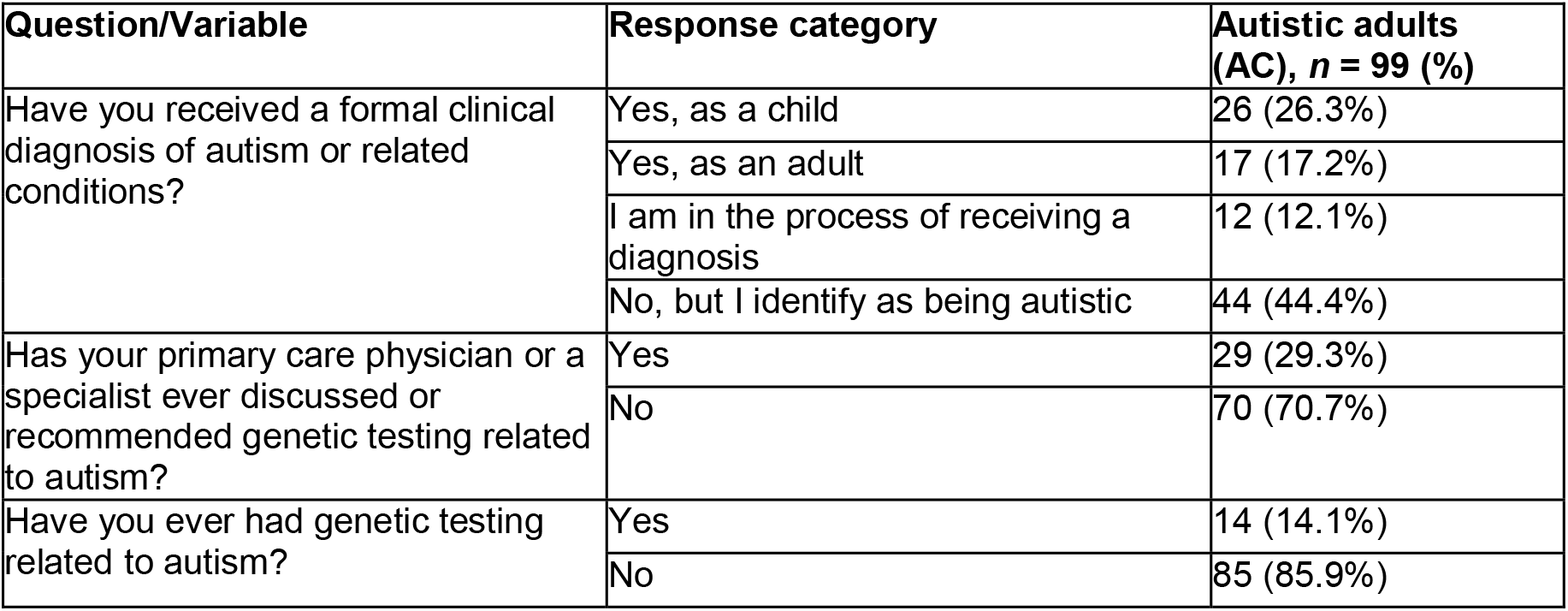
Autistic community additional background questions.

| **Question/Variable** | **Response category** | **Autistic adults (AC), *n* = 99 (%)** |
|---|---|---|
| Have you received a formal clinical diagnosis of autism or related conditions? | Yes, as a child | 26 (26.3%) |
|  | Yes, as an adult | 17 (17.2%) |
|  | I am in the process of receiving a diagnosis | 12 (12.1%) |
|  | No, but I identify as being autistic | 44 (44.4%) |
| Has your primary care physician or a specialist ever discussed or recommended genetic testing related to autism? | Yes | 29 (29.3%) |
|  | No | 70 (70.7%) |
| Have you ever had genetic testing related to autism? | Yes | 14 (14.1%) |
|  | No | 85 (85.9%) |

Among AC participants, 44.4% reported self-identifying as autistic without a formal diagnosis (Table 3). Twenty-nine participants reported that a primary care physician or specialist had discussed autism-related genetic testing with them, and 14 reported having undergone such testing.

### Findings from Genetic Literacy Assessment

Before completing the Education and Assessment of Genetic Literacy (EAGL-long) instrument, participants rated their confidence in their own genetic literacy and familiarity with autism genetics research on a 7-point scale. HCPs reported slightly greater confidence in their genetic literacy than the autistic community panel (mean ± SD, 4.37 ± 1.78 vs. 4.22 ± 1.68) and greater familiarity with autism genetics research (4.04 ± 1.84 vs. 3.50 ± 1.57).

Participants then completed the 46-item EAGL-long instrument, and we compared their scores with those from a previously published representative sample of 2,708 U.S. adults (Barna et al., 2026). The HCP cohort scored significantly higher than the general population on the subjective, applied, situational, and objective genetic literacy domains (all *p* < 0.01), whereas the autistic community panel scored significantly higher on the subjective, applied, and situational domains (all *p* < 0.05) but did not differ from the general population in objective knowledge or genetics skills (*p* = 0.998 and *p* = 0.458, respectively). Most significant differences between the study cohorts and the general population were associated with small effect sizes (Cohen’s *d* = 0.19–0.41), with the exception of objective knowledge in the HCP cohort (*d* = 0.51; Supplemental Tables 2 and 3).

The HCP cohort and autistic community panel performed similarly across four of the five EAGL domains. The HCP cohort scored significantly higher only in objective genetic knowledge (15.44 ± 1.53 vs. 14.50 ± 1.92; Welch’s *t* = 3.84, *p* = 0.0002; Cohen’s *d* = 0.55). The two cohorts did not differ in subjective, applied, situational, or genetics skills domains (all *p* > 0.14).

Associations between recency of genetics/genomics continuing medical education and genetic literacy among healthcare providers are shown in Supplemental Table 4. More recent CME was associated with higher self-rated genetic literacy (ρ = 0.49, FDR-adjusted *p* < 0.001), whereas no significant associations were observed for the EAGL domains after false discovery rate adjustment.

### Participant Evaluation of Intervention Resources

Participants in both cohorts rated all three educational videos favorably for relevance, usefulness, and likelihood of recommendation (Figure 1). Across all three videos, the autistic community panel generally assigned higher ratings for relevance and usefulness than the HCP cohort, whereas HCPs reported a greater likelihood of recommending the videos to colleagues. Mean ratings for each evaluation domain are shown in Figure 1.

**Figure 1:**
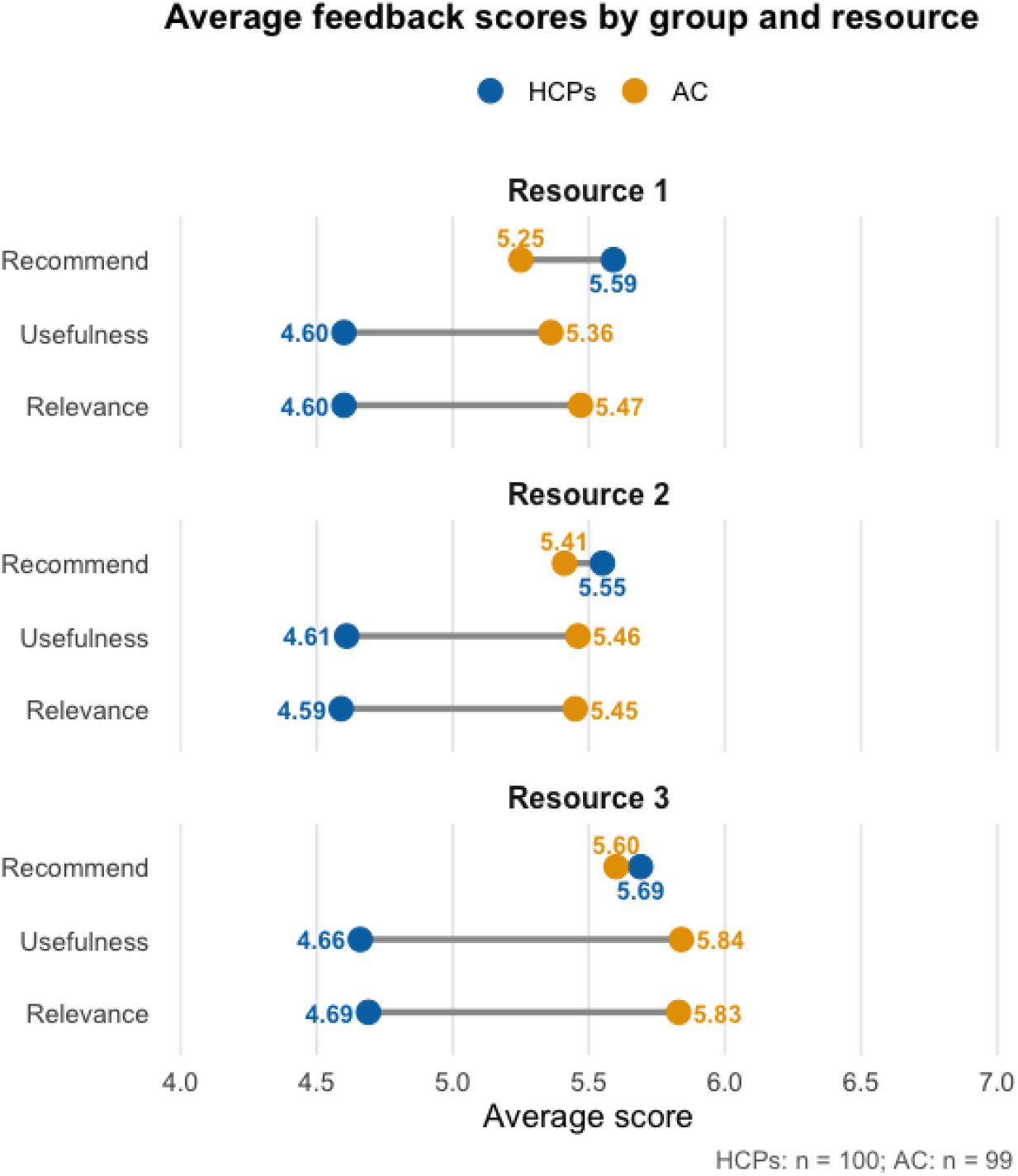
**Average feedback scores for each video, by sample group. Participants used a 1-7 Likert scale but only values 4-7 are represented on the x-axis here for clarity.**

### Impact of the Educational Intervention

Before viewing the educational resources, only 23% of HCPs reported frequently or always discussing genetic testing with autistic individuals or their families (Table 2). Following the intervention, 86% reported that they would be more likely to recommend genetic testing following an autism diagnosis, with 48% selecting the highest category on the 7-point scale. In addition, 88% reported that the videos changed how they would discuss autism and genetics with patients and families.

The intervention also influenced the autistic community panel. Following the intervention, 71% of participants reported that they were more likely to pursue genetic testing after reviewing the educational resources.

### Qualitative Feedback on Intervention Resources

Open-ended responses provided additional context for the quantitative findings. HCP participants were invited to provide optional written feedback after each video. Across the nine open-ended response questions included in the survey (Table 4), the 100 HCP participants submitted 518 comments.

**Table 4.** Quotes from HCPs’ open-response survey feedback questions asked after viewing resources.

| What specific elements of the video 1 did you find useful or not useful to your clinical work? |  |
|---|---|
| Positive Feedback | Negative Feedback |
| “I found all of the information and elements useful for my clinical work because it’s presented in a way that is easy to understand and it is engaging and informative. I find all parts useful especially for someone who doesn’t know a lot about genetics and autism in general. it has a very clear explanation of genetic concepts. I believe this is a valuable source for nurses but not only nurses. I believe it would be a valuable source for everyone in the general population. I would recommend this video to other nurses wanting to learn more about this subject and matter.”<br>Registered Nurse, has referred people for autism diagnostic testing, frequently discusses genetic testing | “While the video was informative, some parts may have been overly simplified, lacking depth in certain areas that require a more detailed explanation. Additionally, some sections contained generalized information that may not be applicable to all clinical specialties.”<br>Psychologist, has diagnosed autistic patient in last 6 months, discusses GT sometimes |
| “The entire information provided in the video is useful to my clinical work. It will help me explain this topic clearly to my patients.”<br>Family Medicine Doctor, has recommended people for autism diagnostic testing, sometimes discusses genetic testing | “The claim that humans share 60% of their DNA with banana is misleading”<br>Pediatrician, involved in diagnostic process in unspecified way, discusses GT sometimes |
| “It provided a simple explanation that could be shown to patients who have no background knowledge of genetics.”<br>Psychiatrist, has referred patients for autism diagnostic testing, discusses GT about half of the time | “its more involved than I need, I don't do the diagnosing”<br>Registered Nurse, occasionally discusses genetic testing |
| “as an RN, I find it helpful to find new ways to teach my patients and other community members about different diagnoses. A lot of the time, I have siblings question why they're different from their siblings, so the whole genetic sequencing section of the video would be super useful”<br>Registered Nurse, has been involved in autism diagnosis process in unspecified way, rarely discusses GT |  |
| “The video provided clear and evidence-based information, which is highly relevant to clinical practice. It included practical demonstrations of techniques that can be directly applied in patient care, making it easier to translate theory into real-world application. Additionally, the explanations were concise and easy to understand, helping reinforce key concepts. The inclusion of case studies and real-life scenarios made the content engaging and applicable to a variety of clinical settings.”<br>Psychologist, has diagnosed a patient with autism in the last 6 months, frequently discussing genetic testing |  |
| What specific elements of the video 2 did you find useful or not useful to your clinical work? |  |
| Positive | Negative |
| “Viewing the genetic components of autism from a different perspective.”<br>Registered Nurse, occasionally discusses genetic testing | “I don't know how this would change my clinical practice. If you have a positive ADOS or<br>other assessment you'll likely send a patient for ABA therapy along with other therapies they may need. If you have suspicions for other intellectual disabilities or disorders you should evaluate for them specifically. For example, ID and large ears<br>or macrochordisim [sic] you might send testing for fragile X. I dont know how useful genetic testing with results of unknown significance is for a patient who you presume has autism.”<br>Pediatrician, diagnosed autism in last 3 years, rarely discusses GT |
| “I think that this helps break things down and makes things clearer it would also be useful for patients with a new diagnosis to watch”<br>Registered Nurse, rarely discusses genetic testing | “to many big words used and to quick of speech rate for the average person to absorb the information”<br>Registered Nurse, involved in autism diagnostic process in unspecified way, frequently discusses GT |
| What specific elements of the video 3 did you find useful or not useful to your clinical work? |  |
| Positive | Negative |
| “It would be a good resource for my patients to watch to understand testing better and to understand cost wait times and be less Afraid of the actual process”<br>Registered Nurse, recommended patients for autism diagnostic testing, rarely discusses GT | “I find it difficult to apply this to my general practice. A lot of my patients do not have the resources to spend \$250 on a test of unknown significance. If my patient has epilepsy that is refractory and would need a genetic test I'd order it. But for a patient with developmental delay or autism I would feel hesitant about ordering a test as a shotgun approach. It's like picking your nose without a plan on what to do with your booger.”<br>Pediatrician, has diagnosed autism in last 3 year, rarely discusses GT |
| “It could be shown to patients and save me some time answering questions”<br>Registered Nurse, recommended seeking autism diagnostic testing, occasionally discuss GT | “Because we don't diagnose patients with autism, and especially discuss things related to price or treatment for those with autism in my role, I didn't find the video useful.”<br>Registered Nurse, never discuss genetic testing |
| Do you have any other comments about the videos? On the material covered by the videos, how the information is presented, etc. |  |
| Positive | Negative |
| “The video was brief, well structured, and easy to understand. The material was clearly defined in straightforward language, supplemented by helpful pictures that reinforced significant points. Complicated genetic vocabulary was clearly explained without seeming overwhelming. Incorporating real-life applications or common situations would even improve it. It was a<br>good presentation in general, informative and enjoyable.”<br>Neuropsychologist, discusses genetic testing sometimes | “I would like the video to go into a bit more detail of the entire process of how all this works. This is very "to make a long story as short as possible" and also to give the viewer ways of remembering all<br>of this complex information better. It's a really great starting point, though, for how complex the information is!”<br>Nursing Assistant, never discusses genetic testing |
| “I wish everyone get the chance to watch this educative video, specially parents who have kids with Autism to understand and stop blaming one another.”<br>Nurse Practitioner, frequently discusses genetic testing | “The information presented in this video is way too high-level for a patient/family member to grasp. It was too high-level for me, as a healthcare professional. I don't think going into so much complex detail is necessary when there isn't any groundwork laid. I feel like this video would just be more overwhelming than helpful to someone who genuinely wanted to learn more about genetic testing. Also, the audio quality is really inconsistent and distracting.”<br>Registered Nurse, not involved in diagnostic process, never discusses GT |
| “Great content! I found it useful since I have a child with autism.”<br>Registered Nurse, involved in diagnostic process in unspecified way, rarely discusses GT | “The script was disjointed, and the tonal difference between the woman and man was noticeable. The HIPAA and GINA slides were too simplistic, and the GINA information should have been on<br>an additional slide, not just spoken.”<br>Psychiatrist, discusses genetic testing sometimes |

Responses were generally positive and indicated that many participants learned information they had not previously encountered, including the diagnostic yield of different genetic testing approaches, current recommendations for autism genetic testing, and the potential clinical benefits of testing. Several participants commented that the resources would improve their ability to discuss genetics and genetic testing with patients and families. Others felt that portions of the material oversimplified autism genetics or covered concepts they already knew, while a small number indicated that the resources were less applicable because their roles did not include diagnosing autism or referring patients for genetic testing.

The autistic community panel was also asked, *“What are your thoughts on genetic testing and how have these videos changed them?ℍ* Ninety-five of the 99 participants responded. Responses reflected a broad range of perspectives regarding autism genetic testing, ranging from strongly supportive to strongly opposed. However, regardless of participants’ initial opinions about testing, many described improvements in their understanding of the testing process and greater confidence discussing genetic testing with healthcare providers or making informed decisions about whether testing was appropriate for themselves or their families. Participants who expressed more favorable views after the intervention frequently attributed this change to learning about the potential clinical benefits of testing, including the identification of co-occurring conditions and improved access to support. Participants who remained skeptical most commonly cited concerns regarding privacy, financial cost, limited perceived clinical utility after an autism diagnosis, or broader ethical concerns surrounding genetic testing.

One participant who identified as autistic described how the videos increased their confidence in an ongoing decision to pursue testing:

> “I feel genetic testing can be very helpful. We are actually looking into it already for my child and these videos made me a little more confident that they can help.”

Another participant emphasized that the resources increased confidence without necessarily changing an underlying opinion about testing:

> “My thoughts haven’t changed much, but it’s good to know more about the process. Being informed is great for improving confidence and awareness when it comes to having testing done.”

Other participants remained skeptical about the value of testing. One participant diagnosed with autism as a child commented:

> “Seems like a waste of money. These videos pretty much confirmed that. I don’t have hundreds of dollars to spend on testing. If it’s able to identify other conditions like epilepsy, then that’s a good thing. But it still seems unnecessary, because I’d probably find out about my epilepsy in another way.”

Additional representative quotations illustrating both supportive and critical perspectives on autism genetic testing are provided in Supplemental Table 5.

## DISCUSSION

This study demonstrates that a brief, video-based educational intervention was well received by both healthcare providers and autistic adults and substantially increased healthcare providers’ reported willingness to discuss and recommend genetic testing following an autism diagnosis. Building on our previous work identifying barriers to provider communication (Wang et al., 2024), this study evaluated a practical educational intervention designed to address those barriers. Although reported intentions do not necessarily translate into changes in clinical practice, the quantitative and qualitative findings together suggest that brief digital educational resources may support more informed discussions between healthcare providers, autistic individuals, and their families (Chen et al., 2024; Cutica et al., 2024; Lewis et al., 2020).

Although the intervention increased participants’ reported willingness to recommend or pursue genetic testing, it is important to note that genetic testing for autism specifically is not appropriate or desirable in every circumstance. There are many reasons why individuals and families may decide not to pursue testing, including ethical concerns, financial barriers, and logistical challenges (Meng et al., 2025; Wills et al., 2025). Likewise, research has shown that even when families express interest in testing, they often report insufficient information to make informed decisions (Rivera-Figueroa et al., 2022), but co-produced educational toolkits have improved the delivery of genetic healthcare in other populations with neurodevelopmental disabilities (Strnadova et al., 2025). Our previous work found that healthcare providers similarly reported limited education and confidence discussing autism genetic testing with patients (Wang et al., 2024). The goal of this intervention was therefore not to encourage universal uptake of genetic testing, but rather to improve communication so that autistic individuals and their families can make informed decisions based on accurate information and current clinical recommendations (Hyman et al., 2020; Schaefer & Mendelsohn, 2013).

Our findings suggest that improving communication, rather than simply increasing genetics knowledge, may represent an important opportunity for improving implementation of current recommendations for autism genetic testing. Following the intervention, most healthcare providers reported that they were more likely to recommend genetic testing and that the videos changed how they would discuss autism genetics with patients and families. Together with the qualitative feedback from both participant groups, these findings suggest that brief educational resources may help address communication barriers identified previously by providers9 while supporting autistic individuals and families in informed discussions about genetic testing with their healthcare providers.

Interestingly, both the HCP and autistic community cohorts demonstrated relatively high genetic literacy compared with the previously published U.S. general population sample (Barna et al., 2026). We reported similar findings in the SPARK autism research community (Little et al., 2022). Despite this relatively high baseline genetic literacy, only 23% of healthcare providers surveyed here reported frequently or always discussing genetic testing before the intervention. In addition, more recent genetics/genomics continuing medical education was associated with higher self-rated genetic literacy but not with objectively measured genetic literacy across the EAGL domains (Supplemental Table 4). Together, these findings suggest that genetics knowledge alone may not be the primary barrier to discussing or recommending autism genetic testing in clinical practice (Wang et al., 2024). Instead, providers may benefit from educational resources that emphasize practical communication strategies in addition to genetics education (Hamilton et al., 2017).

Although the videos were not fully co-produced with autistic community members,34,35 feedback from the community panel suggests that the resources were both acceptable and informative. Following the intervention, 71% of autistic participants reported that they were more likely to pursue genetic testing, while qualitative responses frequently described increased confidence discussing genetic testing with healthcare providers and a better understanding of both the potential benefits and limitations of testing. Importantly, participants who remained hesitant about testing generally cited privacy, cost, or concerns about clinical utility rather than confusion about the information presented. These findings suggest that educational resources can support informed decision-making without assuming that all autistic individuals will reach the same conclusions regarding genetic testing.

The two study cohorts evaluated the educational resources from different perspectives. Autistic participants consistently rated the videos as more relevant and useful than healthcare providers, whereas healthcare providers reported a greater likelihood of recommending the resources (Figure 1). This pattern is likely explained by the different contexts in which the intervention was evaluated (Supplemental Document 1) Healthcare providers considered the videos as resources for clinical practice and communication with patients, whereas autistic participants evaluated their relevance to their own healthcare experiences. Likewise, providers were asked whether they would recommend the videos to colleagues, while autistic participants were asked whether they would recommend them to other autistic individuals or family members.

As noted above, a referral for genetic testing typically cannot occur until a separate, non-referring provider has both diagnosed the patient and independently decided to refer. This handoff has direct implications for medical geneticists’ workflow. Some genetics clinics have already responded by incorporating educational videos directly into the visit itself, nearly doubling clinic throughput and reducing wait times (Barton et al., 2025). Our findings suggest that deploying similar resources further upstream – before or at referral – could extend these efficiency gains to the diagnosing providers and families who precede that visit.

This study has several limitations. The HCP cohort contained a higher proportion of nurses and nursing-related professions than physicians or psychologists, reflecting the composition of the Prolific participant pool rather than the broader workforce involved in autism diagnosis (Autism Speaks, 2025). Nurses play an active role in the diagnostic process, including directing families to free evaluations through early intervention services when autism screenings are positive, or when parents have concerns after a negative screening (Dunlap & Filipek, 2020). Although many participants reported involvement in the autism diagnostic process and discussions of genetic testing, the sample should not be considered representative of U.S. healthcare providers who diagnose autism or routinely order genetic testing. Likewise, the autistic community panel included participants who self-identified as autistic without a formal diagnosis. We chose to retain these participants because self-identification may reflect barriers to obtaining a formal diagnosis, particularly in adulthood (Banker et al., 2025; Newton et al., 2025; Rabba et al., 2025), and because the educational intervention was designed to improve communication for individuals considering or discussing genetic testing regardless of diagnostic pathway.

The educational videos developed for this study are publicly available and could be incorporated into future clinical or research settings, including delivery before genetics or autism-related clinic visits. Future studies should evaluate whether these resources improve patient-provider communication and increase appropriate uptake of genetic testing in routine clinical practice. The substantial increase in providers’ reported willingness to discuss and recommend genetic testing observed here suggests that brief, targeted educational resources may represent an effective strategy for supporting implementation of current clinical recommendations while promoting informed, patient-centered communication about autism genetics.

### AUTHOR CONTRIBUTIONS

**Olivia G. Pilson**: Conceptualization, formal analysis, investigation, methodology, project administration, visualization, writing – original draft, and writing – review and editing. **Lily S. Barna**: Conceptualization, formal analysis, investigation, methodology, and writing – review and editing. **Tyler Grimes**: Formal analysis, visualization, and writing – review and editing. **Taryn A. Salako**: Investigation and writing – review and editing. **Gabriela M. Ramírez Renta**: Investigation, resources, and writing – review and editing. **Amy Wang**: Conceptualization, formal analysis, investigation, methodology, project administration, resources, writing – original draft, and writing – review and editing. **Chris Gunter**: Conceptualization, funding acquisition, investigation, methodology, resources, supervision, writing – original draft, and writing – review and editing

## ACKNOWLEDGMENTS

We thank the Social and Behavioral Research Branch members at NHGRI for suggestions throughout the work. The contributions of the NIH authors are considered Works of the United States Government. The findings and conclusions presented in this paper are those of the authors and do not necessarily reflect the views of the NIH or the US Department of Health and Human Services.

## ETHICS DECLARATION

Both of our research protocols (IRB001103 for the pilot, IRB002400 for larger study) were deemed exempt by the Office of Research Protection of the National Institutes of Health (NIH) and approved by the Institutional Review Board at the National Institutes of Health. All participants provided informed consent for the study. Verified participants of the pilot study received $50 Amazon gift cards, while verified participants in the larger study who completed the survey were provided with a $10 credit through Prolific.

## DATA AVAILABILITY

R code for the tests performed in the paper is available at https://github.com/ChrisG-EMU/Pilson-et-al-PAGES-paper. The data that support the findings of this study are available from the corresponding author upon reasonable request.

## ADDITIONAL INFORMATION

Supplemental Tables 1-5 and Documents 1-3 are available online.

## DECLARATION OF AI AND AI-ASSISTED TECHNOLOGIES IN THE WRITING PROCESS

During the preparation of this work the authors used ChatGPT-5.2 through the CHIRP-NIH Community Chatbot, in order to condense text in the introduction. In addition, we used US HHS-licensed Claude Sonnet 4.2 and 4.6 to co-generate R code for statistical analysis and figures. After using each tool, the authors reviewed and edited the content as needed and take full responsibility for the content of the publication.

